# Estimation of the Potential Impact of COVID-19 Responses on the HIV Epidemic: Analysis using the Goals Model

**DOI:** 10.1101/2020.05.04.20090399

**Authors:** John Stover, Newton Chagoma, Isaac Taramusi, Yu Teng, Rob Glaubius, Guy Mahiane

## Abstract

We applied a simulation model of HIV to analyze the effects of 3 and 6-month disruptions in health services as a result of COVID-19. We found that disruptions to primary prevention programs (male circumcision, behavior change programs, condom distribution) would have small but transitory effects on new infections that might be more than offset by reductions in commercial and multi-partner sex due to lock downs. However, if COVID-19 leads to disruptions in ART services the impacts on mortality could be severe, doubling or tripling the estimated number of HIV deaths in 2020.

## Background

The COVID-19 pandemic has caused serious disruptions to health systems in countries with large numbers of hospitalizations and shortages of protective equipment. Although the impact has been small so far in sub-Saharan Africa it is expected to grow substantially in the coming months. Significant impact on health systems would have consequences in many areas, including HIV. Many countries have already instituted stay-at-home orders that limit mobility and are expected to affect access to health services.

In Zimbabwe, as of April 30, there were 40 cases, 4 deaths and only 5 have fully recovered. The country has put in place measures to prevent and control COVID-19 that include; social distancing (such as banning large gatherings and advising individuals not to socialize outside their households), border closures, school closures, measures to isolate symptomatic individuals and their contacts, and large-scale lockdowns of populations. The country is still under lockdown although there was some relaxation of the policy to the manufacturing and farming industry. The lockdown is indefinite hence the government released guidance for the provision of HIV services as follows: VMMC-limited to walk in clients only at health facility; testing - scale up of self-testing; Pre-exposure prophylaxis (PrEP) - Individuals already on PrEP should be given 3 months prescription and side effects assessment done over social platforms; and ART clients should be given 3-6 months prescription, with priority groups of elderly (50+ years) and adolescent receiving 6 months prescription.

In Malawi, Covid-19 was declared a national disaster on March 20, 2020, and the first Covid-19 case was registered on 2^nd^ April 2020. Consequently, the government developed the National COVID-19 Preparedness and Response Plan, intensified calls for preventive measures, and issued a moratorium on schools, pubs, air transport services except those deemed essential, social gathering to less than 100 people, among others. As a pre-emptive measure, the country also declared a 21-day national lockdown on 14^th^ April, and planned to shutdown central markets, permit only essential travel, restrict public gatherings to less than 10 people, and permit public transport services during the peak hours only and close borders. Other stipulated measures included expedited recruitment of additional 2000 healthcare workers to strengthen the capacity of the health systems, decongest prisons, and close borders. The lockdown was meant to social distance the entire population, and thus break the chain of transmission. However, this has been challenged in the high court arguing that it violated other constitutional provisions – the court stay order was yet to be vacated at the time of this publication. As of 30^th^ April, the country registered a cumulative total of 37 confirmed cases of Covid-19 -- 27 active cases, 3 deaths and 7 recoveries https://malawipublichealth.org/.

While these measures clearly have an impact on flattening the epidemic curve, they also have the potential of disrupting delivery of healthcare services. The HIV program in Malawi has clearly been affected, and among others: suspended provision of voluntary medical male circumcision, PrEP and TB Preventive Therapy, condom distribution to walk in clients, routine and scheduled viral load monitoring for stable patients, teen clubs and other patient support groups. The program has also limited the number of ART clinics days to only once a week and encouraged ART providers to dispense 6-months ARV supply especially for those on first-line regimen, prioritize children failing first-line treatment and incorporate cough and fever screening when providing ART services (MoH-DHA Guidelines on Covid-19).

## Methods

We applied an existing HIV simulation model to examine the potential effects of COVID-19 in 12 countries in sub-Saharan Africa. Goals is an HIV epidemic simulation model that estimates new HIV infections arising from behaviors of distinct risk groups (female sex workers and their male clients, men and women with non-regular partners, faithful couples, men who have sex with men and people who inject drugs). The model includes prevention interventions (both bio-medical and behavioral) and treatment. The model tracks people living with HIV (PLHIV) over time by CD4 count and ART status. Disease progression parameters that determine rates of CD4 decline and mortality of those not on ART are specified by CD4 count. For those on ART, mortality is dependent on age, sex, CD4 count at treatment initiation and duration on treatment [1].

People on ART are tracked by their CD4 count when they start treatment and their time on treatment. The CD4 count of those who drop out of treatment is assumed to revert immediately to the level at which treatment was initiated, similar to findings from studies of treatment interruption that CD4 counts revert quickly to pre-treatment levels [2]. Mortality rates for people living with HIV not on ART are determined by current CD4 count and age, as shown in Table 1. These rates were estimated to match survival rates from HIV cohorts in sub-Saharan Africa before highly active ART was widely available. [3]

**Table 1.** HIV-related annual mortality rates for people living with HIV not on ART

|  | Annual Off-ART HIV Mortality by Age |  |  |  |
| --- | --- | --- | --- | --- |
|  | 15 - 24 | 25 - 34 | 35 - 44 | 45 - 54 |
| > 500 | 0.005 | 0.004 | 0.005 | 0.005 |
| 350 - 500 | 0.011 | 0.01 | 0.013 | 0.013 |
| 250 - 349 | 0.026 | 0.026 | 0.036 | 0.032 |
| 200 - 249 | 0.061 | 0.069 | 0.096 | 0.080 |
| 100 - 199 | 0.139 | 0.185 | 0.258 | 0.203 |
| 50 - 99 | 0.321 | 0.499 | 0.691 | 0.513 |
| < 50 | 0.737 | 1.342 | 1.851 | 1.295 |

We applied the model to 12 countries: Cameroon, Cote d’Ivoire, Eswatini, Kenya, Lesotho, Malawi, Mozambique, Nigeria, South Africa, Tanzania, Uganda and Zimbabwe. Data on the number of men, women and children on ART and voluntary medical male circumcision (VMMC) are from national programs. Behavioral data (condom use, numbers of sexual partners) are drawn from national surveys: Demographic and Health Surveys (DHS), Public Health Impact Assessments (PHIA) in the general population, and Integrated Behavioral and Biological Surveys (IBBS) in key populations. The models were fitted to surveillance and survey data on HIV prevalence.

## Prevention scenarios

For four countries (Malawi, Mozambique, Uganda and Zimbabwe) we examined the effects on HIV incidence and mortality of a three-month disruptions of services due to lockdowns or health system capacity constraints. Service utilization is assumed to return to pre-lockdown levels at the end of the three-month period.

We examined the scenarios listed below. Scenarios were implemented incrementally; for example, the BCC scenario includes VMMC scenario effects on top of interruptions to behavior change interventions.

- **Base**: No impact of COVID-19. Coverage of all interventions except VMMC remains at 2019 levels. VMMC coverage increases according to current trends.
- **VMMC**: No new circumcisions during the lockdown. Circumcisions then resume at their prelockdown rate. The long-term effect is a three-month delay in the trend of increasing circumcision coverage.
- **BCC**: All behavior change interventions are stopped for 3 months. This includes community mobilization, condom distribution, school-based programs and outreach to key populations. The result is an increase in the number of partners and a reduction in condom use.
- **Reduction in Sexual Activity (RedSA)**: During the lockdown there is no casual or commercial sex for heterosexuals or men who have sex with men.
- **PMTCT**: No prevention of mother-to-child transmission (PMTCT) prophylaxis services are offered during the lockdown.
- **ART 3 months**: All ART patients lose access to ART for 3 months.

Separately, for all 12 countries, we modeled the effects of a six-month disruption of ART services on mortality. We assumed that everyone would stop ART for the six months in order to quantify the maximum potential impact.

## Results

The results for primary prevention are shown in Table 2 and Figure 1. The effects of a 3-month shut down of VMMC services are small since most VMMC clients are young and not yet sexually active or in the age group of highest risk. The effects of reductions in behavior change programs raise new infections about 5% in the short term but the effects are small in the longer term. This analysis assumes that behaviors would become risky immediately as the programs to support behavior change are stopped. In reality, people may not revert to riskier behaviors immediately, so a short-term disruption may have little or no effect on behaviors, although the effect could larger if condom supplies are restricted.

**Table 2.** Percent change in new infections and HIV-related deaths due to 3-month lock down for primary prevention and 3- and 6-month lock downs for ART to control COVID-19. Numbers represent the percentage increase in new infections and AIDS deaths relative to the Base scenario.

|  | Change in 2020 |  |  |  | Cumulative Change 2020-2025 |  |  |  |
| --- | --- | --- | --- | --- | --- | --- | --- | --- |
|  | Malawi | Mozambique | Uganda | Zimbabwe | Malawi | Mozambique | Uganda | Zimbabwe |
| **New Infections** |  |  |  |  |  |  |  |  |
| VMMC only | 0% | 0% | 0% | 0% | 0% | 0% | 0% | 0% |
| +BCC | 6% | 6% | 9% | 4% | 1% | 1% | 2% | 1% |
| +Reduced higher risk sex | -8% | -11% | -6% | -4% | -1% | -2% | 0% | 0% |
| +PMTCT | 1% | -6% | 8% | 13% | 0% | -1% | 1% | 2% |
| +ART 3 months | 10% | -2% | 14% | 19% | 3% | -1% | 2% | 5% |
| **AIDS Deaths** |  |  |  |  |  |  |  |  |
| VMMC only | 0% | 0% | 0% | 0% | 0% | 0% | 0% | 0% |
| +BCC | 0% | 0% | 0% | 0% | 0% | 0% | 0% | 0% |
| +Reduced higher risk sex | 0% | 0% | 0% | 0% | 0% | 0% | 0% | 0% |
| +PMTCT | 4% | 3% | 8% | 6% | 1% | 1% | 3% | 2% |
| +ART 3 months | 54% | 40% | 50% | 30% | 17% | 11% | 29% | 21% |

**Figure 1.**
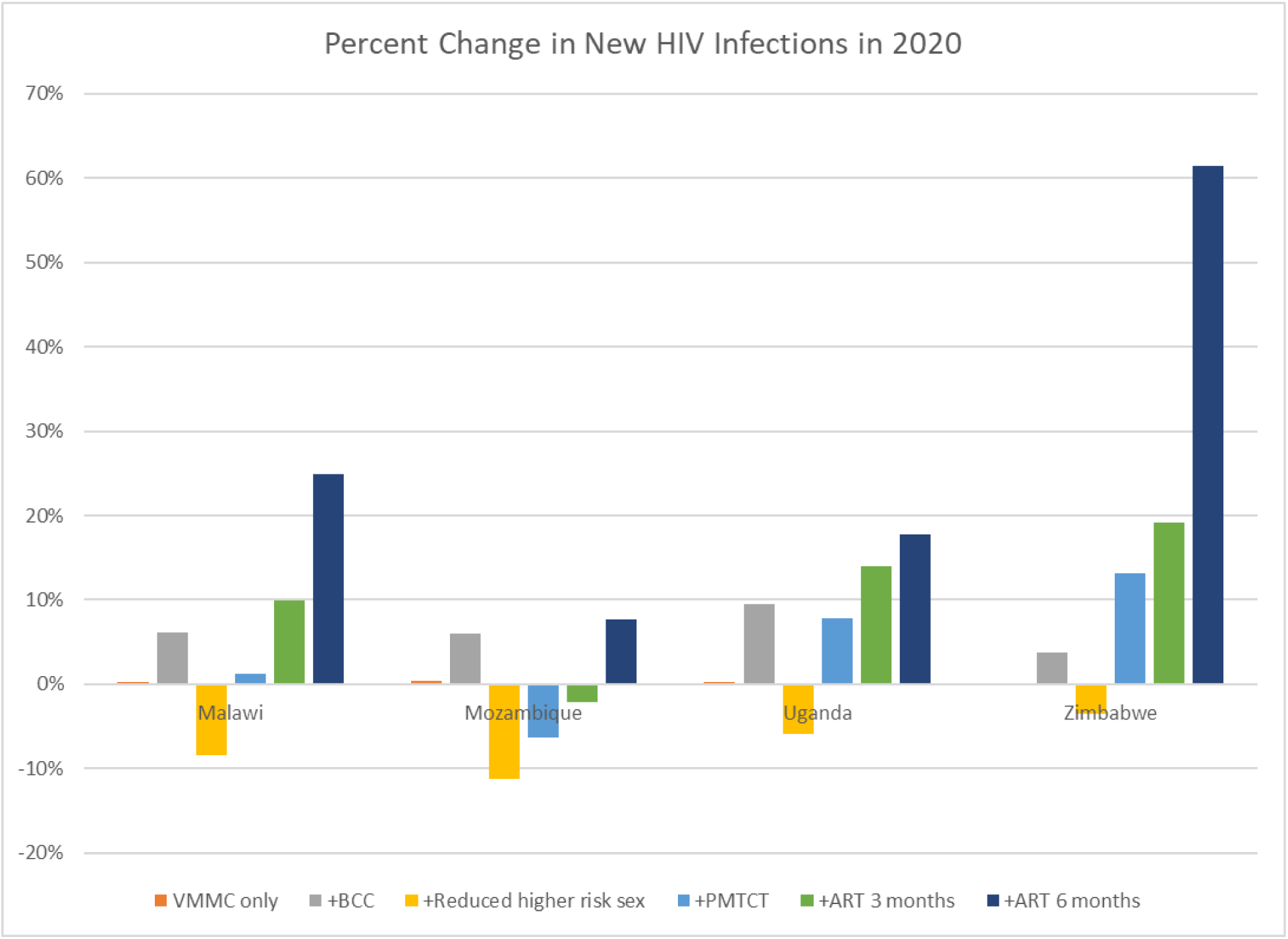
New HIV Infections

If the disruption results in reduced higher risk sex (because bars, clubs and other venues where people meet partners are closed) the result could be fewer new infections (4-11%, the negative bars in Figure 1) in the short-term, but with little long-term effect.

A cessation of PMTCT activities could lead to significant increases in the number of new child infections in 2020. In Figure 2 we show the effects of three scenarios:

- Three-month disruption leading to no women newly starting ART
- Three-month disruption leading to cessation of all prophylaxis among pregnant women
- Six-month disruption leading to cessation of all prophylaxis among pregnant women

**Figure 2.**
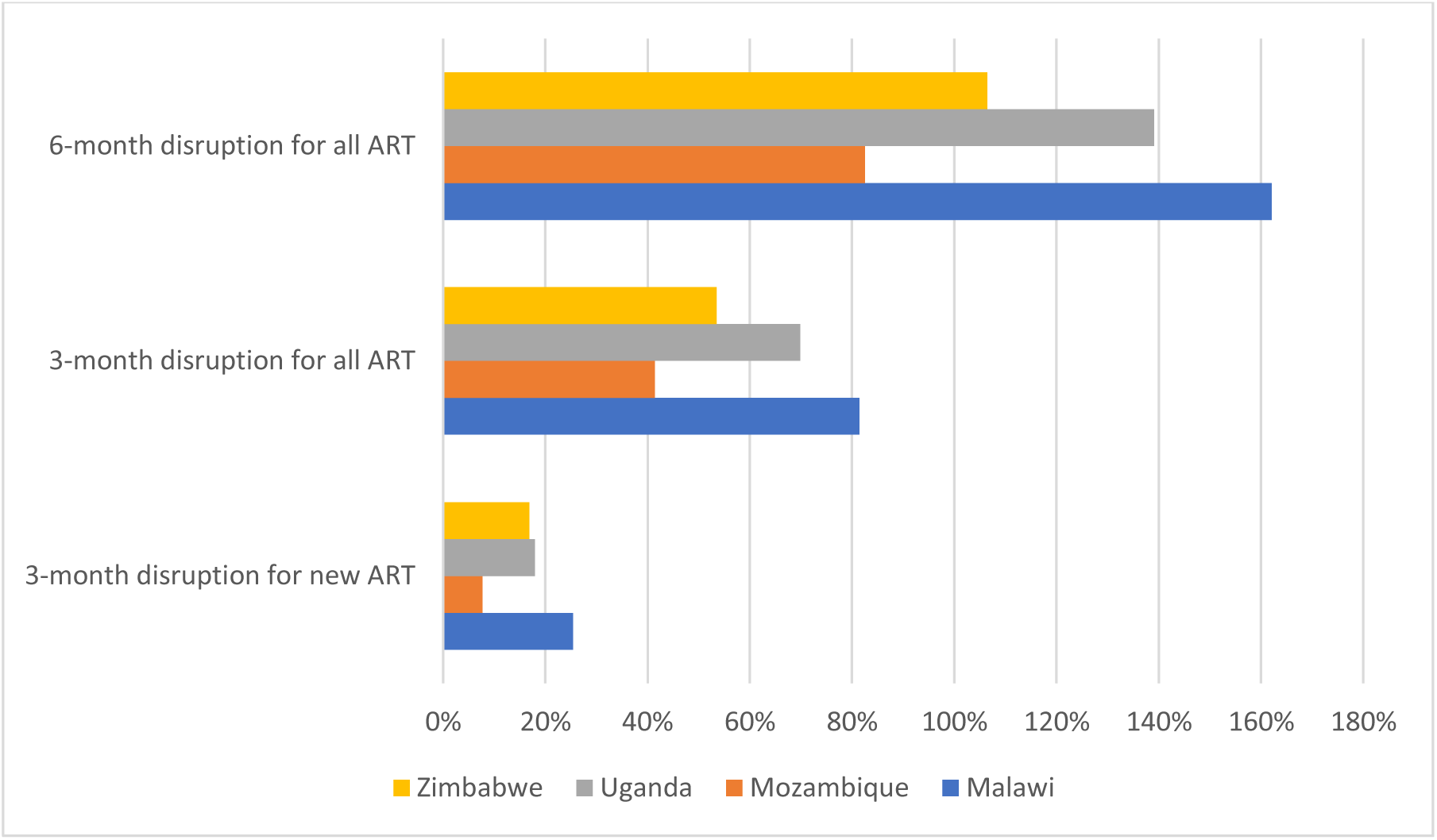
Percentage increase in new child HIV infections

Currently PMTCT coverage is high in most countries resulting in few new child infections. Any disruption in PMTCT services could cause large increases, from about 10% for disruption to new patients, to 100% or more for 6-month disruptions to all PMTCT services.

The effects of ART disruptions will be felt both on new infections and deaths. The number of excess deaths in 2020 and for the longer period 2020-2025 are shown in Table 2. For comparison, Table 2 also shows UNAIDS estimates of HIV-related deaths in 2018 and the number people on ART in 2018. [2]

Countries with larger number of people on ART coverage will generally see more deaths than those with fewer people on ART, but high ART coverage will result in a lower mortality rate since many ART patients have high CD4 counts and, therefore, low mortality even without ART. The percentage increases can easily exceed 100% since currently high ART coverage has led to few HIV deaths. The longer-term impacts are generally less than the short-term impact since some people who would die during the disruptions would have died before 2030 without the disruptions.

**Table 3.** Excess HIV-related deaths from a 6-month disruption of ART

| Country | Estimated HIV-related deaths in 2018 [2] | Number on ART in 2018 | Excess deaths in 2020 due to lock down | Excess deaths 2020-2030 due to lock down in 2020 |
| --- | --- | --- | --- | --- |
| Cameroon | 18,000 | 280,000 | 18,000 | 9,000 |
| Cote d'Ivoire | 16,000 | 250,000 | 13,000 | 12,000 |
| Eswatini | 2,400 | 180,000 | 6,100 | 7,000 |
| Kenya | 25,000 | 1,000,000 | 58,000 | 44,000 |
| Lesotho | 6,000 | 210,000 | 14,000 | 11,100 |
| Malawi | 13,000 | 810,000 | 34,000 | 33,000 |
| Mozambique | 54,000 | 1,200,000 | 92,000 | 88,000 |
| Nigeria | 53,000 | 1,000,000 | 65,000 | 58,000 |
| South Africa | 71,000 | 4,900,000 | 210,000 | 211,000 |
| Tanzania | 25,000 | 1,200,000 | 48,000 | 42,000 |
| Uganda | 23,000 | 940,000 | 52,000 | 97,000 |
| Zimbabwe | 21,000 | 1,200,000 | 48,000 | 59,000 |

## Limitations

We assume that people stopping ART return immediately to the CD4 count they had when the initiated ART. While studies show that CD4 counts decline rapidly after dropping off ART the assumption of immediate return to pre-treatment levels will generally over-estimate the immediate effects on mortality but may be reasonable for lock downs of six-months or more.

Our assumption of no casual or commercial sex during the stay-at-home period may over-estimate the reduction in incidence if some people do not comply with the policy.

The Goals model operates on a time step of 0.1 years but inputs on service coverage are annual, so we approximated the effects of short period of disruption by reducing the annual coverage rate. This will spread out the impact in time but the total effects over one- and five-year periods should be reasonably well estimated.

## Discussion

Our results suggest that the population level effects of COVID-19 on new HIV infections will be small and transitory. The effects could be quite large for individuals who acquire HIV due to interrupted access to PrEP or condoms, but at the population level these effects would be short term.

On the other hand, the effects on mortality of disruptions in ART services could be quite large with at least a doubling of HIV-related deaths and up to three times as many deaths in some cases. The longer-term effects are generally somewhat smaller as the lock down just advances deaths that would have occurred later into 2020.

For a three-month disruption these estimates of impact on new infections are probably overly pessimistic as behavior change may be sustained and ARV supply disruptions may not be total. The assumption of a three-month disruption may be optimistic, however, as some restrictions may stay in place longer and the diversion of resources to fight COVID-19 could restrict essential services for a longer period. In the longer term the effects could be mild, especially if effects to improve health systems to fight COVID-19 have system-wide benefits; or could be more severe than shown here if health systems are severely affected by loss of personnel.

The actual impact of disruptions on ART mortality is likely to be much less than shown here as most programs have starting multi-month dispensing to avoid interruptions in adherence even if they have not yet expanded to all patients. A longer disruption of more than six month would have more severe consequences, if programs cannot offset the impact of service disruptions with alternative approaches, such as home delivery of drugs.

## Disclosures

The authors declare that they have no conflicts of interest. No human subjects or records were involved with this work. Funding was received from the Bill & Melinda Gates Foundation. The funder had no involvement in the research.

## Data Availability

Model projection files are available on request from the authors.

## Notes

### Competing Interest Statement

The authors have declared no competing interest.

### Funding Statement

This work was partially funded by the Bill & Melinda Gates Foundation under grant OPP 1991665

